# Economic Evaluation alongside the Trial of Selective Early Treatment of Patent Ductus Arteriosus with Ibuprofen in Extreme Preterm Babies

**DOI:** 10.1101/2025.06.24.25330184

**Authors:** Chidubem B. Ogwulu, Mark Monahan, Samir Gupta, Nimish V. Subhedar, Jennifer L. Bell, David Field, Ursula Bowler, Elizabeth Hutchison, Sam Johnson, Wilf Kelsall, Justine Pepperell, Sunil Sinha, Kayleigh Stanbury, Jonathan Wyllie, Pollyanna Hardy, Edmund Juszczak, Tracy Roberts, the Baby-OSCAR Collaborative Group

## Abstract

**Objective:** To conduct an economic evaluation alongside the placebo-controlled trial of selective early treatment of patent ductus arteriosus with ibuprofen in extreme preterm babies.

**Design:** Within-trial economic evaluation alongside a multicentre, randomized, masked, placebo-controlled trial.

**Setting:** Thirty-two UK tertiary neonatal intensive care units.

**Sample:** Babies born between 23^+0^ to 28^+6^ weeks’ gestation, less than 72 hours old and confirmed by echocardiography to have a large PDA.

**Methods:** A cost-effectiveness analysis was conducted from the National Health Service (NHS) and NHS and Personal Social Services perspectives. Nonparametric bootstrapping was used to estimate incremental costs and outcomes which were reported as cost-effectiveness acceptability curves.

**Main outcome measures:** Cost per additional major outcome averted.

**Results:** Ibuprofen is less costly with an average cost per participant of £126,465 compared to £133,260 (-£6,808 (95% CI: £-17,154 to £3,537) for placebo. But it is less effective in terms of major outcomes averted with an absolute effect difference of 0.054 (95% CI: -0.014 to 0.121). The differences in mean cost were mainly attributable to the costs of high dependency care (£2,345, 95% CI -£3,435 to £8,126) and intensive care (£6,718, 95% CI - £12,627 to £26,063) which were higher in the placebo arm. The incremental cost-effectiveness ratio was estimated at £126,846 for an additional case of death, or severe or moderate bronchopulmonary dysplasia avoided by 36 weeks of post-menstrual age.

**Conclusions:** Based on the evidence from this trial, ibuprofen would not be recommended. There is insufficient evidence to suggest that any of the observed detrimental impact on effectiveness would be outweighed by the opportunity cost associated of any saving in costs.

## Introduction

Extremely preterm babies are delivered at a gestational age of < 28 weeks [1]. Preterm babies risk serious health problems associated with blood flow, respiration and brain functions. The ductus arteriosus is a blood vessel that allows blood to bypass the lungs to the placenta. This vessel typically closes spontaneously at birth or shortly afterwards. However, in many preterm babies, this vessel remains open - a condition known as Patent Ductus Arteriosus (PDA). In the UK, around 7,000 extremely preterm babies are born annually of which 40% have a PDA that fails to close spontaneously [2].

PDA is associated with several life-threatening short and long-term complications including bronchopulmonary dysplasia (BPD) and a need for prolonged hospitalisation and respiratory support potentially imposing a substantial financial burden on the NHS. The management of a PDA can be conservative (waiting for spontaneous closure), medical or surgical (ligation of the artery).

An early selective treatment approach has been suggested for large PDAs (diameter of ≥1.5 mm). The **O**utcome after **S**elective Early Treatment for **C**losure of Patent Ductus **AR**teriosus in Preterm Babies (Baby-OSCAR) trial [3] was conducted to assess if the selective treatment of large patent ductus arteriosus (PDA) in extremely preterm babies with ibuprofen within 72 hours of birth improves both short-term and long-term health outcomes.

This paper reports the economic evaluation conducted alongside the trial which assessed the cost-effectiveness at two endpoints. The first objective was to assess the relative costs and outcomes of administering Ibuprofen to avoid death and/or moderate or severe bronchopulmonary dysplasia (BPD) (i.e. achieving survival without severe or moderate BPD) at 36 weeks of postmenstrual age (PMA) for extremely preterm babies with PDA. A second objective of the economic evaluation was to assess the cost-effectiveness of the trial intervention in achieving survival at two years of corrected gestational age (CGA).

## Methods

### Trial design and participants

The Baby OSCAR trial is a multicentre, randomized, masked, placebo-controlled trial. Details of the trial are published elsewhere [4]. Briefly, between July 2015 and December 2020, 653 preterm babies with echocardiogram evidence of a large PDA were recruited from 32 tertiary neonatal intensive care units (NICU) across the UK. Written informed consent was provided by the parents of the participants.

Ethical approval was obtained from the UK Medicines and Healthcare Products Regulatory Agency, the NHS Health Research Authority, and the East Midlands–Nottingham 2 Research Ethics Committee. The study was funded by the UK NIHR HTA Programme (HTA 11/92/15). The funder was not involved in the identification, design, conduct, and reporting of the analysis. The inclusion and exclusion criteria are available in *Appendix 1*.

### Intervention and outcomes

The trial medication was three doses of Ibuprofen administered as an intravenous (IV) injection. The trial had two clinical endpoints – (i) an initial short-term endpoint of either 36 weeks postmenstrual age (PMA) or discharge from the initial hospital stay, and (ii) a final (long-term) endpoint of 2 years of corrected gestational age (corrected for prematurity).

### Economic Evaluation

Several short-term secondary clinical outcomes were assessed in the trial, and these are presented in a disaggregated format as part of a preliminary cost–consequence analysis (CCA). The main economic evaluation took the form of a within-trial cost-effectiveness analysis (CEA) performed from the perspective of the NHS/Personal Social Service (PSS) [5]. Thus, only direct healthcare costs were included. The main analysis is based on the primary short-term outcome of the trial which is a composite outcome of deaths avoided and/or cases of moderate or severe bronchopulmonary dysplasia eliminated, at 36 weeks of postmenstrual age. The results are expressed in terms of incremental cost per additional major outcome averted (MOA) where a major outcome is defined as death and/or moderate or severe BPD avoided by 36 weeks of PMA.

A secondary CEA based on the trial’s long-term endpoint and reported in terms of cost per survival without severe or moderate neurodevelopmental disability assessed at two years CGA.

The reporting of the economic evaluation is in keeping with the Consolidated Health Economic Evaluation Reporting Standards (CHEERS) statement [6]. Given the initial short-term endpoint of the trial, neither costs nor outcomes were discounted, however, discounting was applied for the final endpoint at an annual rate of 3.5% [5]. All statistical analyses were performed using STATA version 17 (StataCorp, 2021) [7]. No health economics analysis plan was developed for the economic evaluation.

### Data Collection

Data on both resource use and outcome were collected prospectively during the trial using trial-specific case report forms (CRFs). Information also included data from both an internal pilot phase and the main trial. Data included an initial hospital stay, and by the initial trial endpoint (36 weeks or earlier) babies were either discharged or transferred to another hospital. In addition, for the secondary analysis, health service use until two years, which included the use of primary care services and hospital readmission was collected through a parent-completed questionnaire. Details of the resource use are available in *Appendix 2*.

Unit costs for each resource item (*Table 1*) were identified from several established national sources [8,9] and all costs were expressed in 2022-23 UK pounds Sterling values. Medication costs were extracted from the *British National Formulary for Children* [10] and some costs were also obtained from previous literature [11,12] and from the NHS blood and transplant price list 2020/21 [13]. Cost estimates from earlier years were inflated to 2022-23 prices using the Hospital and Community Health Services (HCHS) pay and prices index [8]. In cases where there are different categories associated with resource use, weighted averages were used.

**Table 1:** Unit cost of resource use items (2022-23 prices)

| Resource use items | Unit cost (£) * | HRG code | *Source/year |
| --- | --- | --- | --- |
| **Trial Medication** |  |  |  |
| Ibuprofen (Pedea) per course | 288 | N/a | BNFC 2023 [10] |
| **Short-term outcomes** |  |  |  |
| Blood (packed cell) transfusion | 145 | N/a | NHS Blood and transplant price list 2023/24 [13], NICE, 2015, JPAC, 2015 |
| Sepsis | 5,688 | WJ06D-F | NHS 2021/22 [9] |
| Coagulopathy – (fresh frozen plasma transfusion) | 130 | N/a | NHS Blood and transplant price list 2023/24[13], NICE, 2015[20], JPAC, 2015[21] |
| Exchange transfusion | 1301 | SA42Z | NHS Cost Schedule 2021/22[9] |
| Cerebral Ultrasound | 84 | RD42Z | NHS Cost Schedule 2021/22[9] |
| Intraventricular haemorrhage (IVH) Grade I or 2 | 1,292 | N/a | Tahir et al., 2020[11] |
| IVH Grade 3 or 4 | 1,681 | N/a | Tahir et al., 2020 [11] |
| Hydrocephalus | 3,384 | YQ42Z | NHS Cost Schedule 2021/22[9] |
| Cystic Periventricular Leukomalacia (PVL) | 3,246 | PR06A-B | NHS Cost Schedule 2021/22[9] |
| Non-cystic PVL | 1,562 | PR06C | NHS Cost Schedule 2021/22[9] |
| Gastrointestinal bleeding | 1,741 | FD03A-H | NHS Cost Schedule 2021/22[9] |
| Intestinal perforation | 10,734 | FF23C-E; FF35C-D | NHS Cost Schedule 2021/22[9] |
| Pneumothorax | 2,447 | DZ26G-P | NHS Cost Schedule 2021/22[9] |
| Seizure (Standard EEG monitoring plus a course of IV phenobarbitone) | 825 (700 + 125) | AA81Z | NHS Cost Schedule 2021/22[9], BNFC 2023 [10] |
| Steroids for Chronic lung disease | 24 | N/a | BNFC 2023 [10] |
| ECHO | 116 | RD51C | NHS Cost Schedule 2021/22[9] |
| **Initial trial end baby outcomes** |  |  |  |
| PDA treatment with NSAIDs | 1,275 | N/a | Tahir et al., 2020[11] |
| Surgical ligation of PDA | 2,683 | N/a | Modi et al., 2019[12] |
| Retinopathy of prematurity (ROP) treatment | 1,085 | PP64A-B | NHS Cost Schedule 2021/22[9] |
| ROP Screening | 148 | N/a | Modi et al., 2019[12] |
| Significant Pulmonary haemorrhage | 1937 | PD65A-B | NHS Cost Schedule 2021/22[9] |
| Pulmonary hypertension | 990 | EB15C | NHS Cost Schedule 2021/22[9] |
| NEC Bell stage II and above (surgery) | 5364 | FD01A-E | NHS Cost Schedule 2021/22[9] |
| Non-invasive ventilation | 2267 | DZ27H; DZ27J-K | NHS Cost Schedule 2013/14[22] |
| Low flow Oxygen | 1244 | DZ27L | NHS Cost Schedule 2013/14[22] |
| Invasive ventilation by an endotracheal tube | 4111 | DZ27G | NHS Cost Schedule 2013/14[22] |
| Inotropic support | 6 | N/a | BNFC 2023 [10] |
| Level 1- Intensive care (ICU) | 1868 | XA01Z | NHS Cost Schedule 2021/22[9] |
| Level 2 – High dependency care (HDU) | 1264 | XA02Z | NHS Cost Schedule 2021/22[9] |
| Level 3 – Special care (SCU) | 675 | XA03Z;XA04Z | NHS Cost Schedule 2021/22[9] |
| **Outcomes at 2 years** |  |  |  |
| **Hospital/Community Services** |  |  |  |
| Surgical ligation of PDA | 2,683 | N/a | Modi et al., 2019 [12] |
| Hospital admissions (nights) | 427 | PX56A to PX57C (nights) | NHS Cost Schedule 2021/22[9] |
| Intensive care (nights) | 1,868 | XA01Z | NHS Cost Schedule 2021/22[9] |
| **Outpatient department/clinic** |  |  |  |
| Accident & Emergency | 148 | SC180 | NHS Cost Schedule 2021/22[9] |
| Audiology/Hearing | 281 | SC254 | NHS Cost Schedule 2021/22[9] |
| Eye/vision | 130 | SC216 | NHS Cost Schedule 2021/22[9] |
| General Medical | 156 | SC300 | NHS Cost Schedule 2021/22[9] |
| Paediatric/Neonatal follow up clinic | 293 | SC420 | NHS Cost Schedule 2021/22[9] |
| **Community professionals** |  |  |  |
| Community nurse | 40 | N/a | Jones & Burns 2022[8] |
| Community paediatrician | 360 | SC290 | NHS Cost Schedule 2021/22[9] |
| Dietician | 80 | A03 | NHS Cost Schedule 2021/22[9] |
| Physiotherapist | 139 | A08C1; A08CG | NHS Cost Schedule 2021/22[9] |
| Speech & language therapist | 145 | A13C1; A13CG | NHS Cost Schedule 2021/22[9] |
| Others | 161 | N/a | Jones & Burns 2022[8] |

Resource use was valued using ‘top-down’ costing approaches[14] where aggregated costs at each organisational cost are centred down to units of activity. The trial medication was three doses of Ibuprofen administered as an intravenous (IV) injection and the cost was £72 per 2ml single-use ampoule of Pedea.[10]. The initial dose was 2ml/kg (10mg/kg) followed by two doses of 1ml/kg (5 mg/kg) at 24 and 48 hours. For the trial, it was assumed that a complete dose of the trial medication cost £288 (£144 +£72 +£72). The excesses from the 2nd and 3rd doses were typically discarded. In addition, some babies weighed more than a kilogram, thus a 4th ampoule may have been used for the loading dose (*Appendix 2*).

### Missing Data

Multivariate regressions and t-tests were used to explore whether missingness for costs and outcomes could be predicted by other variables in the existing data [15]. Data were assumed to be missing completely at random if the associations between variables were not statistically significant at the 5% level. Based on this assumption, missing costs were imputed using multiple imputations [16], by applying chained equations with predictive mean matching across 60 imputations .

### Analysis

Prior to the CEA, a cost consequences analysis (CCA) was carried out. A CCA is typically the first step in an economic evaluation in which disaggregated costs and outcomes are presented in their natural units, to see if there is any strategy that shows clear dominance [17].

The total cost during the trial period was estimated by multiplying the resource items used, by the corresponding unit cost followed by summation. We calculated the mean total costs and mean total resource use for the babies across trial arms. To address the skewness inherent in most cost data and the concern of economic analyses with mean costs, the bias-corrected and accelerated (BCa) nonparametric bootstrap methods were used to estimate 95% confidence intervals (CIs) around mean differences by analysing 1,000 resamples [18]. For the nonparametric bootstrap method repeated random samples of equal size as the original sample are drawn with replacements from the data. The mean is calculated from each resample and these bootstrap estimates of the original statistic are then used to build up an empirical distribution for the mean [19].

### Sensitivity Analysis

Sensitivity analysis was used to quantify the inherent uncertainty relating to assumptions and sampling variations in the methods used and ultimately to assess the generalisability of the results. To assess heterogeneity in the trial population, multivariate cost analyses were conducted using seemingly unrelated regressions [23,24], an accepted method that allows for a correlation in the error terms between costs and outcomes [25] and is robust to skewed data. Model covariates included the size of PDA at randomisation; gestational age at birth; age at randomisation; sex; multiple births; mode of respiratory support at randomisation; receiving inotropes at time of randomisation; and centre ID. The selection of covariates was informed by the prognostic variables used by the clinical team. All results were presented as mean values with standard deviation (SD) and where applicable, as mean differences in costs and effects with 95% CIs.

The Incremental Cost Effectiveness Ratios (ICER) was calculated by dividing the difference in mean total cost between the trial arms by the difference in the number of outcomes avoided.

To quantify the uncertainty, which is often attributable to sampling variations, the joint distribution in the mean cost and outcome difference were resampled using non-parametric bootstrapping [26]. This generated 5,000 paired estimates of incremental costs and outcomes which were plotted in a cost-effectiveness plane [27]. A cost-effectiveness plane is a four-quadrant plane which illustrates bootstrap estimates. Depending on the location of the cost-effectiveness result in the quadrant, an intervention may be judged as more effective and more costly (northeast quadrant), more effective and less costly (southeast), less effective and less costly (southwest) or less effective and more costly (northwest) than the alternative intervention.

A cost-effectiveness acceptability curve (CEAC) was constructed to show the probability the use of IV Ibuprofen for the selective treatment of PDA in pre-term babies is a cost-effective intervention compared with placebo across a range of values representing the decision maker’s willingness to pay (WTP) for an additional benefit [5].

Deterministic sensitivity analysis comprised an exploration of alternative unit costs applied to the different resources used or the variation of some parameters while leaving others at their baseline value. When appropriate, these analyses were combined with the stochastic sensitivity analysis.

- *Complete dose of trial medication*: For the trial, all participants were to receive three doses of the trial intervention. For the primary analysis, we estimated the costs based on the actual number of doses received. For this analysis, it was assumed that all participants received all doses.
- *Follow-up costs*: Multiple imputations were performed to impute the missing costs for primary care and resource use data up to two years follow-up. Costs were imputed at the total cost level and the imputation model used 60 imputations.

## Results

### Participants

The results of the Baby-OSCAR trial are reported in detail elsewhere [4]. Six hundred and fifty-three preterm babies were recruited and randomised to either the ibuprofen arm (n=326) or the placebo arm (n=327). Seven babies were excluded from the analysis due to consent withdrawal by the parents to use data that were already collected. Hence, the initial economic analysis, based on the short-term outcome, involved 646 participants: 324 in the ibuprofen arm and 322 in the placebo arm.

### Outcomes

The details of the major outcomes of the trial are presented in *Table S1*. At 36 weeks postmenstrual age or discharge from the hospital, (44 preterm babies in the ibuprofen arm and 33 in the placebo arm) had died or had moderate to severe bronchopulmonary dysplasia (176 vs 169). Thus, major outcomes were averted for 98 (30.8%) babies in the ibuprofen arm and 116 (36.5%) babies in the placebo arm. The absolute effect difference was -0.054 (95% confidence interval -0.121 to 0.014). At two years of age (corrected for prematurity), the major outcomes were averted for 109 (48.4%) babies in the ibuprofen arm and 117 (49.6%) babies in the placebo arm with an absolute difference of -0.020 (95% confidence interval -0.107 to 0.068).

### Cost-consequence Analysis

A breakdown of the resource use data by the trial arms is presented in *Table S2*. Mean healthcare costs per participant by trial arm are presented in *Table S3*. Based on findings from the cranial ultrasound, babies in the Ibuprofen arm, on average, had more intracranial abnormalities than babies in the placebo arm. In contrast, on average, babies in the placebo arm spent more days in intensive care (ITU), high-dependency care (HTU) and special care. Consequently, the length of the initial hospital stay was longer in the placebo arm. There was very little variation in resource use during the initial hospital stay for the majority of variables.

For the two years of follow-up, the most intensive resources were recorded for the PDA operation, visits to the paediatrician and overnight hospital stays and these resources were all higher in the placebo arm than the intervention arm (*Table S2*). The average cost of the trial medication was estimated to be £270 (95% CI £265 to £276) (Table S3).The most substantial costs accrued during the trial by participants during the initial trial end, were for neonatal intensive care unit (ITU) stay, with a mean cost of £50,764 (SD £38,583) per baby for the Ibuprofen arm and £52,898 (SD £50,508) per baby for the placebo arm.

In keeping with the resource use, variables associated with hospital stay were consistently lower in the ibuprofen arm. Thus, the placebo arm had higher costs of £2,101 (95% CI - £4,213 to £8,415); £2,345 (95% CI -£3,435 to £8,126) and £953 (95% CI -£981 to £2,886) for ITU, high dependency unit (HDU) and special care unit respectively (*Table S3*) compared to the ibuprofen arm. For all endpoints by the initial trial end, the biggest cost difference was in nights of admission in the HDU; this was £2,345 (95% CI -£3,435 to £8,126) more in the placebo arm. During the follow-up period, the biggest cost difference was for admissions to the ITU which was higher in the placebo arm compared to the ibuprofen arm with a cost difference of £6,718 (95% CI -£12,627 to £26,063).

The mean total costs by the trial arm for different variables are presented in *Table S4*. For the initial trial endpoint (36 weeks PMA or discharge from hospital), the total cost was £126,465 and £133,260 in the Ibuprofen and placebo arms respectively. Hence this cost was higher in the placebo arm with a mean difference of £6,727 (95% CI: -£3,690 to £17,144) per baby. When the costs for the two-year follow-up are included, the total cost of the trial remained higher in the placebo arm (£137,903) than in the ibuprofen arm (£129,867) with a mean difference of £7,904 (95% CI: -£3,115 to £18,924).

### Primary Analysis

The primary cost-effectiveness outcome based on the short-term primary outcome of the trial was the cost per additional case of death or severe or moderate bronchopulmonary dysplasia (BPD) avoided by 36 weeks of PMA. The ICER which combines the differences in costs in both arms is presented in *Table 2*. The ibuprofen intervention was less costly than placebo, with a cost-saving of £6,809 (95% CI: -£3,537 to £17,154) per baby. However, the intervention was slightly less effective than placebo, with an absolute effect difference of 0.054 (95% CI: -0.013 to 0.120). This means that fewer infants achieved the principal outcome of survival at 36 weeks of postmenstrual age (PMA) without moderate or severe BPD in the ibuprofen arm compared to the placebo arm (98 vs 116 respectively).

**Table 2.** Deterministic base case results.

| Treatment | Total cost (£) | Total effect | Adjusted Cost difference (£)<br>(95% CI) | Effect difference | ICER (£) |
| --- | --- | --- | --- | --- | --- |
| Short-term primary outcome |  |  |  |  |  |
| Ibuprofen | 126,465 | 0.308 | -6,809<br>(-17,154 to 3,537) | -0.054<br>(-0.120 to 0.013) | 126,846 |
| Placebo | 133,260 | 0.365 |  |  |  |
| Two-year follow up primary outcome |  |  |  |  |  |
| Ibuprofen | 129,867 | 0.484 | -11,796<br>(-25,251 to 1,658) | -0.0197<br>(-0.104 to 0.065) | 600,264 |
| Placebo | 137,903 | 0.496 |  |  |  |

Given the differences in costs and effects, the point ICER estimate for Ibuprofen compared to placebo was calculated at £126,846 per additional case of death, or severe or moderate BPD avoided by 36 weeks of PMA.

### Deterministic Sensitivity Analyses

We conducted sensitivity analyses in which we explored the impact of receiving a complete dose of the trial medication (*Table S5*). This did not impact the resulting ICER. For the two-year follow-up data, multiple imputations were used to impute missing costs by applying chained equations with predictive mean matching across 60 imputations (Rubin, 2004).

### Sensitivity Analyses

The result of 5,000 bootstrap replications plotted on the cost-effectiveness plane for the primary analysis is presented in *Figure S1*. Each point on the plane depicts a pair of incremental cost and incremental effectiveness estimates for the comparison between Ibuprofen and placebo. The majority of the scatter plots are in the south-west quadrant. This suggests that the Ibuprofen intervention is less costly but also less effective.

The cost-effectiveness acceptability curve (CEAC) which shows the probability of Ibuprofen being cost-effective at various values of decision-makers’ willingness to pay is presented in *Figure S2*. The probability that Ibuprofen is cost-effective falls as the willingness to pay amount increase. The probability that Ibuprofen is cost effective is less than 40% at the arbitrary £20,000 threshold.

The cost-effectiveness plane and the CEAC for the trial endpoint (two-years CGA) are provided in Figures S3 and S4 respectively. Similar to the initial end-point, the majority of the scatter plots are in the southwest quadrant (*Figure S3*), further suggesting that the Ibuprofen intervention remains less costly but also less effective. However, the probability of the intervention being cost-effective at potentially acceptable WTP thresholds (*Figure S4*) was slightly higher (just over 40%) at the final trial end than at the initial trial end (less than 40%).

## Discussion

### Principal Findings

The cost-effectiveness of ibuprofen in achieving survival at 36 weeks of postmenstrual age (PMA) without severe or moderate bronchopulmonary dysplasia (BPD) for extremely preterm babies with patent ductus arteriosus was estimated. The results suggest that ibuprofen is less costly with an average cost per participant of £126,465 compared to £133,260 (-£6,808 (95% CI: £-17,154 to £3,537) for placebo. But it is less effective with an absolute effect difference of 0.054 (95% CI: -0.014 to 0.121). The differences in cost were mainly attributable to the costs of high dependency care (£2,345, 95% CI -£3,435 to £8,126) and intensive care (£6,718, 95% CI -£12,627 to £26,063) which were higher in the placebo arm. The ICER was estimated at £126,846 for an additional case of death, or severe or moderate BPD avoided by 36 weeks of PMA.

When the uncertainty around all the estimates is incorporated into the analysis, the findings suggest that despite being a potentially cost-saving intervention, the ibuprofen intervention would not be considered cost-effective because of the detrimental impact on infants. The cost-effectiveness plane (Figure S1) which incorporates the uncertainty around each point estimate in the results shows, that relative to placebo, ibuprofen is less effective (with the scatter plot being predominantly to the left of the origin in Figure S1). The cost-effectiveness acceptability curve (CEAC) (Figure S2) suggests that the probability of ibuprofen being cost-effective is less than 50% for all willingness to pay (WTP) values and the probability of cost-effectiveness clearly decreases as the WTP increases.

Following initial discharge, there are other hospital resources as well as primary care resources that could be accessed by the infants due to the trial’s disease focus (PDA), such as neurodevelopmental disability and chronic respiratory problems. In addition, there could be broader societal consequences such as productivity loss, due to both time off work and the lost earnings of parents of children with disabilities [28,29]. Although the trial collected resource and outcome data up to two years of corrected age, these only included primary care services use and did not explore the societal impact. It is of note that whilst the costs of initial hospital care were lower in those allocated to the ibuprofen arm compared to those that were given placebo up to the initial outcome, there is the possibility that if resource use data were available up to the trial final endpoint, these costs could change. However, additional analysis of data from the final endpoint showed that the costs did not change, showing that there is no lasting benefit or late positive effect associated with Ibuprofen.

The economic analyses were shown to be robust as evidenced by the sensitivity analyses, which did not show any significant impact on the primary ICER value.

### Strengths and limitations of the economic analyses

The strength of the CEA is that it is the first economic evaluation alongside a randomised controlled trial that compared the cost-effectiveness of parenteral Ibuprofen for the treatment of PDA in preterm infants. The outcome and resource use data were prospectively collected at different points in the trial using CRF. Unit costs were obtained from standard and recognised sources. In cases where HRGs did not clearly depict our variables, we liaised with the clinical team to decide on the most appropriate HRG. The CEA also benefitted from the robustness of the main analyses and sensitivity analyses.

In terms of limitations, there was apparent sparse use of health service resources in the follow up period to two years which did not necessarily indicate missing-ness but lack of use of health care resources but there is some uncertainty about this. A longer follow up period may reveal additional cost and consequences of the disabilities/impairments present and highlight the extent of degree of differentiation in health service use between the two groups, which would be informative. Our analyses required some pragmatic assumptions regarding the blood volumes and medicines usage. Nonetheless, our assumptions are supported by existing literature and guidelines, and the relatively low-cost impact is unlikely to have any significant impact on the results.

### Comparison with the literature

This is the first UK based RCT to investigate the cost-effectiveness of intravenous ibuprofen in achieving survival at 36 weeks of postmenstrual age (PMA) without severe or moderate bronchopulmonary dysplasia (BPD) for extremely preterm babies with PDA. A recent cohort-based study [30] in Qatar compared the cost-effectiveness of oral versus intravenous ibuprofen treatment in preterm infants with PDA. The authors reported that oral ibuprofen was less costly US $13,356 (95% CI US $13,014–$13,699) and more effective (risk ratio = 0.56; 95% CI, 0.32–0.97; *P* = 0.04) than intravenous ibuprofen [30]. However, a cohort study does not represent the gold standard level of evidence as produced by the randomised controlled trial upon which this study is based.

### Implications for policy

The results of the CEA showed that utilising IV ibuprofen is less costly but less effective. In some cases, in health economic evaluation, it can be acceptable to recommend an intervention that is slightly less effective if the resource savings are such that they can be utilised to better effect elsewhere. However, this is rarely the case for interventions to prevent disabilities in newborn babies – as at this time point there is insufficient evidence to support the likelihood that the detrimental impact on effectiveness is outweighed by the opportunity cost associated with the saving [11]. In this case too, in terms of cost-effectiveness, ibuprofen has less than a 50% probability of being cost-effective at most willingness to pay thresholds, in the primary analysis. Furthermore, should a decision-maker be willing to pay £20,000 to increase the chances of an additional survival at 36 weeks without severe or moderate dysplasia, the probability decreases to less than 30%.

## Data Availability

Data sharing requests will be considered by the BABY OSCAR management group upon written request. After approval of proposal, or with a signed data access agreement investigator involvement will depend on the discussion and agreement that is reached for the use of the data.

