## Appendices for "Economic Evaluation alongside the Trial of Selective Early Treatment of Patent Ductus Arteriosus with Ibuprofen in Extreme Preterm Babies"

### Appendix 1: Inclusion and Exclusion Criteria

**Inclusion criteria**

- Born at 23+ 0–28+ 6 weeks’ gestation
- Less than 72 hours old
- Confirmed by echocardiography to have a large PDA which
- is at least 1.5 mm in diameter (determined by gain optimised colour Doppler),

*And*

- has unrestrictive pulsatile (left to right) flow (ratio of flow velocity in PDA Maximum (Vmax) to Minimum (Vmin) > 2:1)) or, growing flow pattern (< 30% right to left), and no clinical concerns of pulmonary hypertension

In addition:

- The responsible clinician is uncertain about whether the baby might benefit from treatment to close the PDA
- Written informed consent is obtained from the parent(s).

**Exclusion criteria**

Babies will be excluded from participation in the trial if they have:

- No realistic prospect of survival
- Severe congenital anomaly
- Clinical or echocardiography suspicion of congenital structural heart disease that contraindicates treatment with ibuprofen
- Other conditions that would contraindicate the use of ibuprofen (active bleeding especially intracranial or gastrointestinal bleeding, coagulopathy, thrombocytopenia (platelet count < 50,000), renal failure, life threatening infection, pulmonary hypertension, known or suspected necrotising enterocolitis (NEC))
- Indomethacin, ibuprofen, or paracetamol administration after birth

### Appendix 2: Details of Resource Use

The CRFs recorded details on resource use data on various categories and clinical time points.

1. *Post Randomisation* - The number of doses of the trial medication
2. *Randomisation to seven days after the final dose of the trial medication* - Foreseeable serious adverse events (SAE)
3. *Three weeks (18 to 24 days) of age* - Echocardiogram (ECHO) results
4. *36 weeks postmenstrual age or at discharge* - pre-specified baby outcomes at the hospital
5. *Two years Corrected Gestational Age (CGA)* - Health service use.

The CRFs also captured information surrounding open treatments of PDA and the occurrence of necrotising enterocolitis (NEC).

#### Trial Medication (Post Randomisation)

The trial medication was three doses of Ibuprofen administered as an intravenous (IV) injection. The initial dose was 2ml/kg (10mg/kg) followed by two doses of 1ml/kg (5 mg/kg) at 24 and 48 hours**.** The cost of IV Ibuprofen was obtained from the *British National Formulary for Children* (BNFC) [10] as £72 per 2ml single-use ampoule of Pedea.

For the trial, it was assumed that a complete dose of the trial medication cost £288 (£144 +£72 +£72). This is because excesses from the 2^nd^ and 3^rd^ doses were typically discarded. In addition, some babies weighed more than a kilogram, thus the 4^th^ ampoule may have been used for the loading dose.

#### Foreseeable Serious Adverse Events (*Randomisation to seven days)*

Following randomisation, data were collected on foreseeable serious adverse events (SAEs) for each baby up to seven days after the administration of the last dose of the trial medication. Foreseeable SAEs are defined as those events expected in the patient population or as a result of the routine care of a patient.

The resource data on SAEs included information on:

- anaemia for which a blood transfusion was required.
- hyperbilirubinemia that led to an exchange blood transfusion and
- coagulopathy that required treatment. The routine treatment for coagulopathy includes the administration of fresh frozen plasma, packed red blood cells and medications like Vitamin K.

The costs of blood transfusion products such as red blood cells and plasma were obtained from the *NHS Blood and Transplant price list* [13] and applied to the volume of use for each product.

Blood transfusion cost includes the blood product, staff time, disposable, storage wastage and laboratory tests [20]. The cost of neonatal red cells is provided on the NHS price list as £69 per unit [13]. Based on the Joint United Kingdom (UK) Blood Transfusion and Tissue Transplantation Services Professional Advisory Committee (JPAC) guideline for blood transfusion in pre-term babies with anaemia it was assumed that a unit of blood is equivalent to the maximum recommended volume of 10 to 20mls/kg per transfusion [21]. In line with the National Clinical Guideline on Blood Transfusion (NICE, 2015) an additional cost of £76 was added for each unit to cover staff time, disposable and wastage and we estimated a total cost of £145 (£69 + £76) per unit of blood transfused.

A unit of fresh frozen plasma for neonates costs £38 [13]. The total cost for treatment of coagulopathy was estimated as £130 per unit by using an average of the cost of plasma and packed red blood cells plus £76 for staff time.

The trial also collected data on SAEs such as on the incidence of sepsis, gastrointestinal bleeding, intestinal perforation, pneumothorax, hypotension and seizure, and if the participants received any treatment for these events. The costs (Table 1) for managing some of the SAEs were obtained from the NHS Reference Cost Schedule [31].

Medication costs for the management of the SAEs (Table 1) were estimated based on the doses and the frequencies reported on the trial’s CRFs. The management of seizures involves the use of electroencephalogram (EEG) monitoring and the administration of Phenobarbitone as a first-line drug. The trial data provided this information as seizures that required treatment. Hence, this was calculated as the cost of standard long-term EEG monitoring (£700) plus a course of IV phenobarbital (£125). In the event of hypotension, babies were treated with inotropes such as Dobutamine or Dopamine. Steroids were administered to babies with chronic lung disease; this included a total dose of 0.89 mg/kg dexamethasone given over 10 days. These costs were obtained from the BNFC [10]. For babies that needed a cerebral (cranial) ultrasound, information was collected on the types and management of abnormalities identified by the scan.

#### Echocardiogram Results (Three weeks (18 to 24 days) of age)

At around three weeks of age, all babies in both arms had an echocardiogram (ECHO), hence this cost was not included in the analysis. However, there were additional ECHOs conducted for babies that were eligible for open treatment of PDA and these costs were included. The cost of an ECHO was obtained from the NHS Reference Costs [31].

#### Baby Outcome (36 weeks postmenstrual age or at discharge)

For the trial, the primary outcome was assessed at 36 weeks postmenstrual age (PMA) or discharge if this was before 36 weeks. Information was collected on the length of time on respiratory support. Respiratory support could be invasive ventilation - via an endotracheal tube or non-invasive such as nasal continuous positive airway pressure (nasal CPAP), humified high flow nasal cannula therapy (HHFNC) or nasal ventilation. These costs were obtained from the NHS cost schedule 2013/14 [22] and updated to 2022/23 prices.

Open-label treatment of PDA was provided to babies that met pre-set trial criteria. Information was collected on the type of PDA treatment – surgical or medical. Medical treatment involved the use of cox inhibitors including Ibuprofen, indomethacin or paracetamol. The medications were costed based on dosage and frequency. Surgical treatment involves a ligation of the PDA and the costs were obtained from an NIHR Journals paper on national resource for neonates research programmes [12].

Data were also collected on the management of retinopathy of prematurity (ROP), cardiovascular and gastrointestinal complications. Resource use also included the number of nights in the intensive care unit (ITU), high dependency care unit (HDU) and special care unit. The costs for these resources were obtained from the NHS costs schedule [31].

### Resource use for outcomes at two-year follow-up

#### Health service use at two years *(Two years CGA)*

1. The parent questionnaires captured information on any respiratory illness following discharge. Data were collected on the attendance of hospital outpatient clinics such as Accident and Emergency, Audiology, Eye, General Medical and Neonatal/Paediatric follow up. The questionnaires also captured data on primary care service use at two years. These included the number of visits by the community nurse, community paediatrician, dietician, physiotherapist, speech and language therapist and other community services. To cost each primary care resource use, we used the recommended average duration of each category [8].

#### Unforeseeable Serious adverse events

Data on SAEs were collected using SAE forms. Only clinically specified SAEs deemed to have arisen from the trial Intervention were considered to be relevant to the economic analysis.

### Appendix 3 List of the investigators in the Baby OSCAR Collaborative Group

Saulius Satas, Margaret Connon, Stephen Main, Susan MacFarlane, Anthony Wilfred Ross Kelsall, Katherine Bradly Russell, Helen Shelley, Beth Berthlecon, Sajeev Job, Anand Kamalanathan, Sharon Hughes, Lucy Lewis, Aung Soe, Jaideep Singh, Eve Irvine, Katie Price, Laura Thrasyvoulou, Juneka Begum, Jacqueline Daglish, Vishna Rasiah, Anju Singh, Rachel Jackson, Efygenia Kotsia, Amy Woodhead, Abby Twiss, Maxine Heather Barrow, Elizabeth Simcox, Sam Wallis, Rachel Wane, Savithiri Sivashankar, Emily Andrews, Heather Collier, Chi-Ning Gerrard, Caroline Cowman, Bev Hammond, Frances Pickering, Robert John Madar, Sarah-Jane Sharman, Alison Stolton, Jonathan Wyllie, Caroline Buckley, Amanda Forster, Helena Smith, Suzanne Bell, Porus Bustani, Pauline Bayliss, Rachel Sellars, Lynne Smart, Liz Taylor, Beth Lally, Lawrence Miall, Nicola Balatoni, Suzanne Laing, Collette Spencer, Sarah Thornton, Lindsay Uryn, Laura Dalton, Katherine Pettinger, Charlotte Reilly, Jonathan Cusack, Marie Hubbard, Rosalind Astles, Maria Sharpe, Jennifer Smith, Nimish Subhedar, Karen Harvey, Joanne Windrow, Patrick McGowan, Amy Beasley, Sateeshkumar Somisetty, Yvonne Millar, Olaitan Adesiyan, Jenny Baker, Santosh Pattnayak, Helen Harizaj, Aimee Harris, Sarah Jones, Alison Youdale, Rahul Roy, Samantha Claire, Supriya Bhoomaiah, Karen Few, Katherine Lloyd, Amy Nichols, Laura Playne, Jay Banerjee, Batia Gourin, Zoe McClure, Kirupalini Mariampillai, Sundar Satyamurthy, Christopher Kissack, Sally Yip, Lynn Clark, Bharathi Rao, Eileen Killen, Jennifer McGowan, Muriel Millar, Mary O'Neill, Angela Abbate, Rachel Anderson, Julie Brown, Patrick Lawlor, Judith Ratcliffe, Eileen Rogers, Akaolisa Egbeama, Joanna Lees, Claire Lodge, Natalie Morgan, Raju Narasimhan, Paula Sugden, Sundeep Harigopal, Julie Groombridge, Tracey Downes, Donovan Duffy, Naomi Hayward, Anay Kulkarni, Izabela Andrzejewska, Vania Oliveira, Sundar Sathiyamurthy, Arindam Mukherjee, Nicola Booth, Karen Dockery, Clare Jennings, Louise Weaver-Lowe, Katherine Birchall, Majid Abu-Harb, Natalie Talbot, Paul Corrigan, Siddhartha Sen, Alison Davies, Angela Harris, Ajay Sinha, MaySze Chang, Caroline Francia, Ivone Lancoma-Malcolm, Gail Falder, Rainer Ebel, Mrinalini Rajimwale, Francesca Brewer, Rebecca Grenfell, Nicola Watts, Laura Wild, Nicolas Aldridge, Susan Dale, Jo Gmerek, Kerri McGowan, Samir Gupta, Sundaram Janakiraman, Alex Ramshaw, Wendy Cheadle, Chidambara Harikumar, Nazakat Merchant, Shabana Malik, Suminthra Naidu, Rona Verdadero, Amit Gupta, Shermi George, Claire Moloney, Vimal Vasu, Gopala Krishnan, Denise Vigni, Jennifer Bell, Ursula Bowler, Christina Cole, Kerrianne Dempster, Clare Edwards, Pollyanna Hardy, Nina Jamieson, Edmund Juszczak, Ann Kennedy, Andy King, Marketa Laube, Louise Linsell, David Murray, Heather O'Connor, Charles Roehr, Kayleigh Stanbury, Julia Sutton, Joy Wiles, Tracy Roberts, Chidubem Okeke Ogwulu, Jane Greenaway, Pauline Shephard, Volker Straub, Justin Carter, Ben Snook, Denis Azzopardi, Michael Weindling, Narender Aladangady, Sophie Welch, Tim Clayton, Alan Montgomery, David Edwards, Heike Rabe

### Appendix Figures

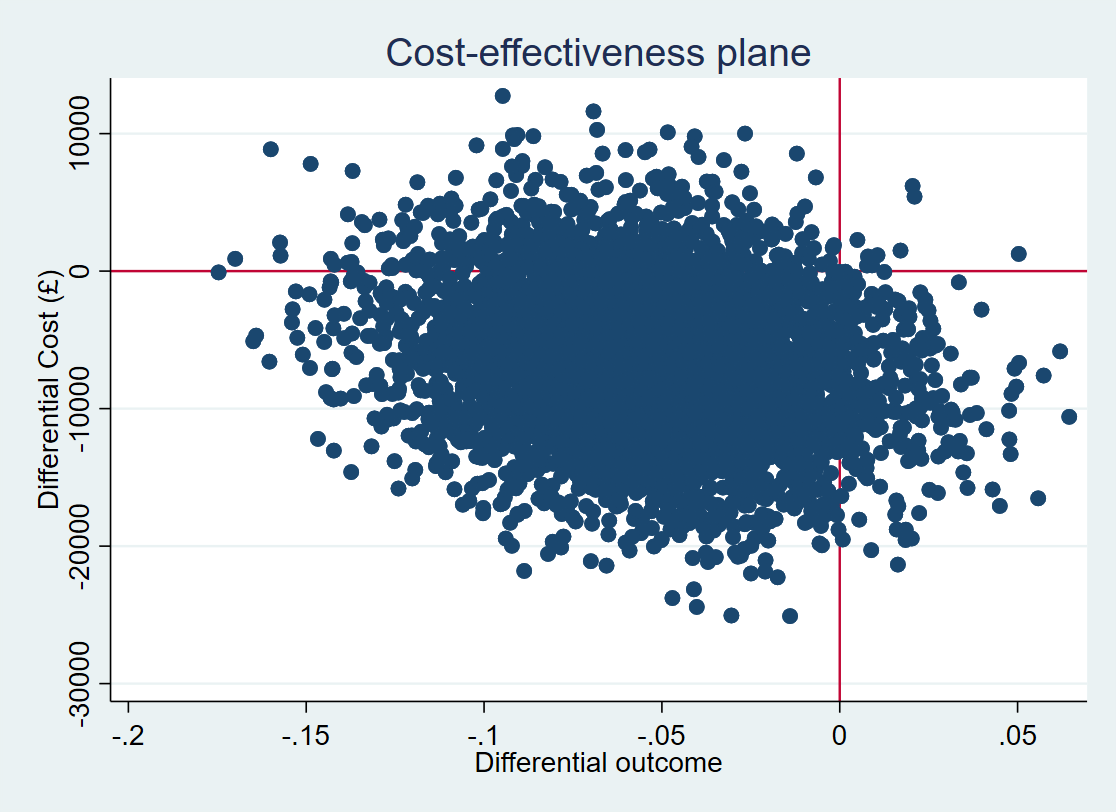

##### Figure S1. Cost-effectiveness plane for the Primary analysis based on the trial's short-term primary outcome.

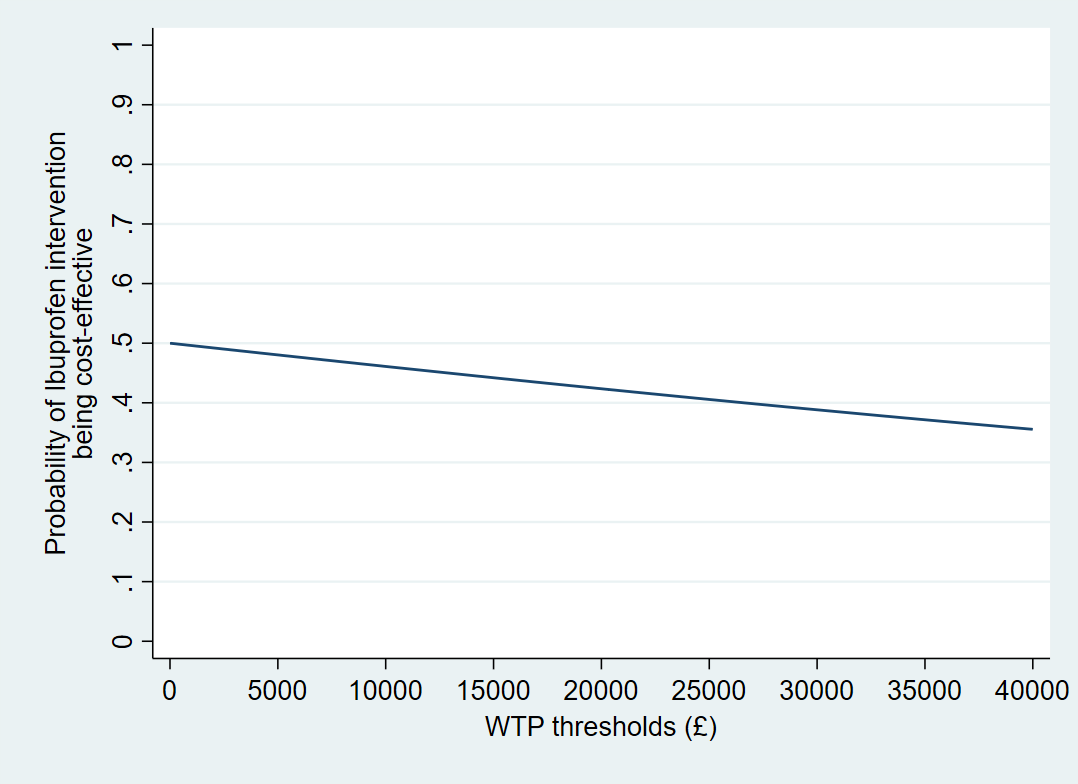

##### Figure S2. CEAC for the Primary analysis based on the trial's short-term primary outcome

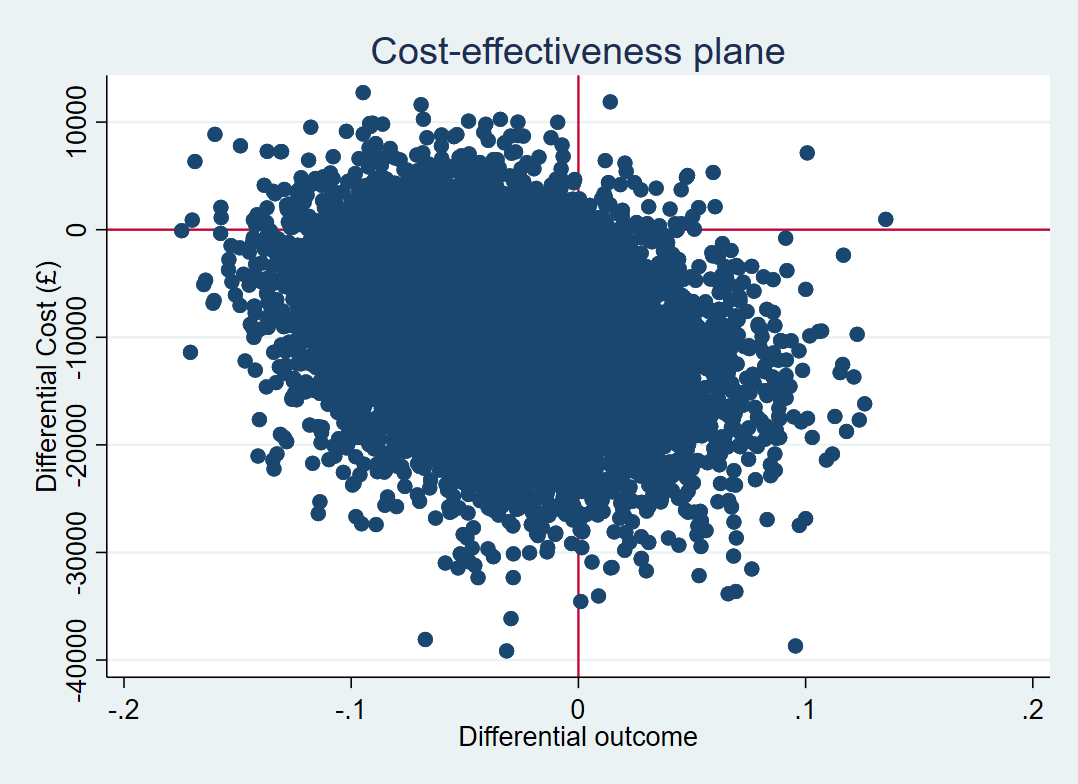

##### Figure S3. Cost-effectiveness plane for the Primary analysis based on the trial's end primary outcome.

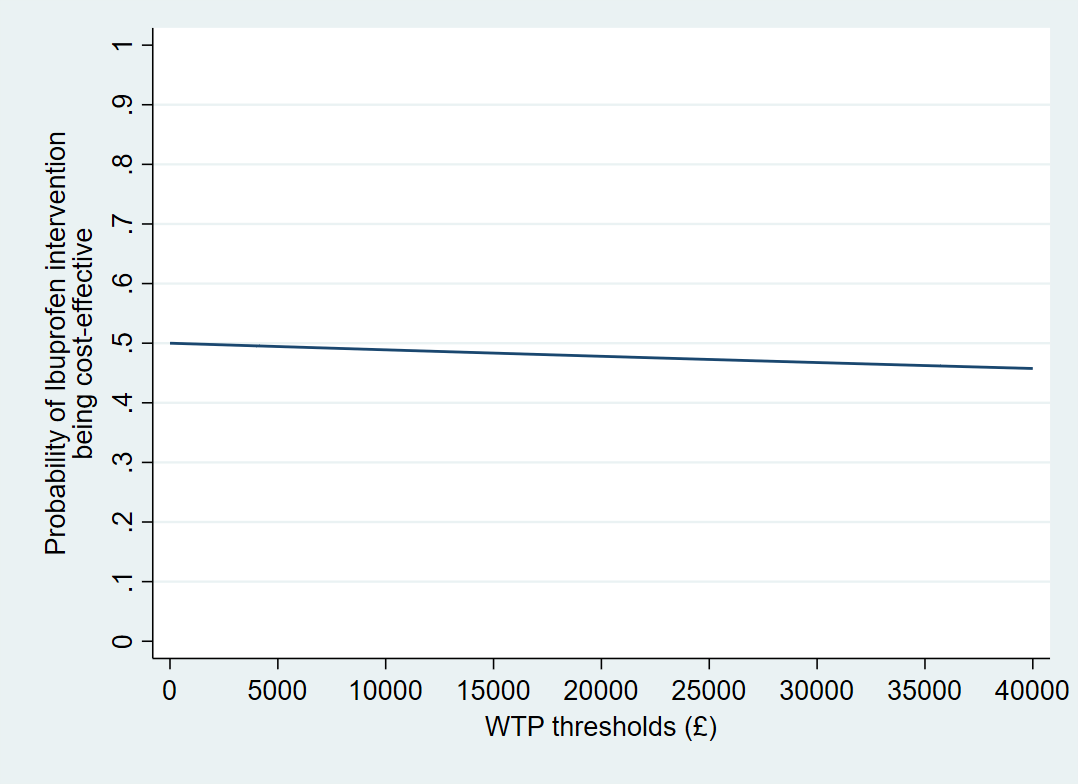

##### Figure S4. CEAC for the Primary analysis based on the trial's end primary outcome

Appendix Tables

##### Table S1. Trial Outcomes

| **Outcomes** | **Ibuprofen N=324** | | **Placebo N=322** | | **Bootstrap difference (95% CI)** | | |
| --- | --- | --- | --- | --- | --- | --- | --- |
|  | **n/N (%)** | **Mean (SD)** | **n/N (%)** | **Mean (SD)** | **Adjusted mean** | **Lower CI** | **Higher CI** |
| ***Primary short-term outcome*** : | | | | | | | |
| Major outcome averted# | 98/318 (30.82) | 0.308 (0.462) | 116/318 (36.48) | 0.365 (0.482) | -0.054 | -0.121 | 0.014 |
| ***Primary short-term outcome components*** | | | | | | | |
| Deaths averted *(survival at hospital discharge)* | 280/324  (86.42) | 0.864  (0.343) | 289/322 (89.75) | 0.898  (0.304) | -0.029 | -0.077 | 0.017 |
| No Moderate to severe BDP | 98/274 (64.2) | 0.642 (0.480) | 116/285 (59.3) | 0.593 (0.492) | 0.052 | -0.018 | 0.123 |
| ***Primary outcome at two years:*** | | | | | | | |
| Major outcome averted* | 109/225 (48.44) | 0.484 (0.501) | 117/236 (49.58) | 0.496 (0.501) | -0.020 | -0.107 | 0.068 |
| # Primary short-term outcome was defined as a major outcome averted (MOA) where a major outcome is defined as death and/or moderate or severe BPD avoided by 36 weeks of PMA.  * Primary outcome at two years was defined as a survival without severe or moderate neurodevelopmental disability assessed at two years corrected gestational age.  BDP = bronchopulmonary dysplasia | | | | | | | |

##### Table S2. Disaggregated Resource Use

| **Resource items** | **Ibuprofen N=324** | | | **Placebo N=322** | | | **Bootstrap difference (95% CI)** | | |
| --- | --- | --- | --- | --- | --- | --- | --- | --- | --- |
|  | **N** | **Mean** | **SD** | **n** | **Mean** | **SD** | **Adjusted Mean difference** | **Lower CI** | **Higher CI** |
| **Trial medication** | | | | | | | | | |
| Average number of doses received | 324 | 2.778 | 0.639 | 322 | 2.804 | 0.628 | -0.025 | -0.122 | -0.073 |
| ***Secondary binary short-term outcomes (Mean/SE)*** | | | | | | | | | |
| Sepsis treatment | 306 | 0.121 | 0.327 | 303 | 0.119 | 0.324 | 0.001 | -0.051 | 0.054 |
| Coagulopathy | 320 | 0.097 | 0.296 | 320 | 0.084 | 0.278 | 0.011 | -0.033 | 0.055 |
| IVH_1_2 | 324 | 0.284 | 0.452 | 322 | 0.304 | 0.461 | -0.023 | -0.092 | 0.046 |
| IVH_3_4 | 324 | 0.139 | 0.346 | 322 | 0.106 | 0.308 | 0.032 | -0.017 | 0.081 |
| Hydrocephalus | 324 | 0.040 | 0.197 | 322 | 0.028 | 0.165 | 0.012 | -0.016 | 0.041 |
| Cystic PVL | 324 | 0.046 | 0.210 | 322 | 0.028 | 0.165 | 0.018 | -0.012 | 0.047 |
| Non-cystic PVL | 324 | 0.006 | 0.078 | 322 | 0.006 | 0.079 | 0.0002 | -0.012 | 0.012 |
| Gastrointestinal bleeding | 324 | 0.028 | 0.165 | 322 | 0.028 | 0.165 | -0.002 | -0.027 | 0.027 |
| Intestinal perforation | 25 | 0.88 | 0.332 | 23 | 0.957 | 0.209 | -0.056 | -0.199 | 0.086 |
| Pneumothorax | 313 | 0.013 | 0.113 | 310 | 0.016 | 0.126 | -0.004 | -0.021 | 0.014 |
| Seizure | 313 | 0.022 | 0.148 | 311 | 0.016 | 0.126 | 0.006 | -0.015 | 0.027 |
| PDA medical treatment | 324 | 0.133 | 0.340 | 322 | 0.0255 | 0.436 | -0.118 | -0.177 | -0.060 |
| PDA Surgical treatment | 324 | 0.028 | 0.165 | 322 | 0.096 | 0.295 | -0.069 | -0.105 | -0.033 |
| ROP screening | 324 | 1.435 | 0.917 | 322 | 1.519 | 1.128 | -0.086 | -0.242 | 0.071 |
| ROP treatment | 324 | 0.139 | 0.346 | 322 | 0.140 | 0.347 | -0.004 | -0.055 | 0.047 |
| Steroids | 323 | 0.263 | 0.441 | 322 | 0.255 | 0.436 | 0.006 | -0.055 | 0.068 |
| Pulmonary haemorrhage | 322 | 0.075 | 0.263 | 322 | 0.056 | 0.230 | 0.019 | -0.019 | 0.056 |
| Diuretics | 324 | 0.293 | 0.456 | 322 | 0.388 | 0.488 | -0.093 | -0.161 | -0.024 |
| Pulmonary hypertension | 324 | 0.053 | 0.223 | 321 | 0.050 | 0.218 | 0.002 | -0.032 | 0.036 |
| NEC requiring surgery | 323 | 0.059 | 0.236 | 322 | 0.078 | 0.268 | -0.020 | -0.058 | 0.018 |
| ***Secondary short-term outcomes (Mean/SD)*** | | | | | | | | | |
| Volume of transfusions | 190 | 31.16 | 14.40 | 179 | 31.82 | 14.74 | -0.60 | -3.26 | 2.06 |
| Exchange blood transfusion | 0 | - | - | 1 | 1 | - | - | - | - |
| Cerebral ultrasounds | 324 | 5.90 | 3.80 | 322 | 5.82 | 3.25 | 0.05 | -0.44 | 0.56 |
| ECHO | 320 | 3.438 | 2.146 | 318 | 3.774 | 2.134 | -0.33 | -0.63 | -0.03 |
| Invasive ventilation (number) | 324 | 0.941 | 0.235 | 322 | 0.966 | 0.182 | -0.026 | -0.058 | -0.006 |
| Invasive ventilation (days) | 324 | 17.910 | 18.656 | 322 | 18.643 | 20.032 | -0.921 | -3.290 | 1.448 |
| Non-invasive ventilation (number) | 324 | 0.892 | 0.311 | 322 | 0.941 | 0.236 | -0.046 | -0.086 | -0.006 |
| Non-invasive ventilation (days) | 324 | 41.660 | 30.614 | 322 | 43.037 | 36.184 | -1.314 | -6.303 | 3.675 |
| Low flow Oxygen (number) | 324 | 0.784 | 0.412 | 322 | 0.807 | 0.395 | -0.019 | -0.083 | -0.044 |
| Low flow oxygen (days) | 324 | 20.019 | 18.482 | 322 | 20.615 | 21.877 | -0.418 | -3.644 | 2.809 |
| Neonatal intensive care days | 313 | 27.176 | 20.655 | 311 | 28.318 | 27.038 | -1.125 | -4.426 | 2.176 |
| Neonatal high dependency care days | 307 | 37.958 | 29.850 | 309 | 40.003 | 29.471 | -1.855 | -6.313 | 2.602 |
| Neonatal special care | 307 | 24.378 | 17.216 | 307 | 25.909 | 18.811 | -1.411 | -4.233 | 1.410 |
| Days of Inotropic support | 133 | 4.857 | 5.235 | 120 | 3.85 | 4.216 | 0.847 | -0.241 | 1.936 |
| Courses of antibiotics | 195 | 2.221 | 1.992 | 197 | 2.274 | 1.809 | -0.065 | -0.430 | 0.301 |
| **Long-term outcomes at 2 years** | | | | | | | | | |
| PDA Operation | 150 | -0.0067 | 0.3928 | 164 | 0.01829 | 0.3028 | -0.0304 | -0.1165 | 0.0557 |
| In Night admission | 74 | 8.5811 | 10.3209 | 84 | 9.7143 | 14.0615 | -1.4218 | -5.09997 | 2.256342 |
| Night ITU | 8 | 8.5000 | 7.4066 | 16 | 10.625 | 10.1053 | -3.72201 | -13.99135 | 6.547328 |
| Visit to the Emergency | 46 | 3.0652 | 3.0214 | 70 | 4.1143 | 5.1907 | -0.927738 | -2.503186 | 0.6477098 |
| Auditory visit | 44 | 3.4545 | 5.1690 | 43 | 2.7209 | 3.0026 | 0.0324944 | -1.36986 | 1.434848 |
| Eye visit | 66 | 4.0000 | 4.0038 | 72 | 3.6389 | 3.5017 | 0.3789439 | -0.896768 | 1.654656 |
| General medicine visit | 24 | 3.7500 | 1.6746 | 26 | 5.8846 | 9.4798 | -0.7307358 | -4.017983 | 2.556511 |
| Paediatrics/Neonatal follow-up | 96 | 4.7292 | 3.2686 | 115 | 5.6696 | 6.2796 | -0.8287573 | -2.131498 | 0.4739832 |
| Community paediatrician visit | 30 | 9.7667 | 27.9824 | 22 | 4.4545 | 2.9233 | 6.604135 | -6.173811 | 19.38208 |
| Other hospital visit | 34 | 4.7353 | 4.6795 | 56 | 6.2321 | 6.7366 | -1.534533 | -4.03839 | 0.9693229 |
| Community nurse visit | 55 | 15.6727 | 26.0214 | 71 | 11.1268 | 13.7902 | 4.9705 | -3.6205 | 13.5615 |
| Dietician visit | 46 | 4.2826 | 4.2981 | 57 | 5.1579 | 5.7501 | -0.9276723 | -3.047406 | 1.192061 |
| Physiotherapist visit | 64 | 8.8906 | 17.1289 | 66 | 8.3485 | 13.8147 | 1.487111 | -4.311839 | 7.28606 |
| Speech therapist visit | 38 | 4.5526 | 6.3489 | 38 | 5.1579 | 6.8321 | -0.8590572 | -3.89591 | 2.177795 |
| Other professional visit | 19 | 5.0526 | 4.6245 | 16 | 12.8125 | 23.2529 | -1.006068 | -8.956065 | 6.943928 |

*The follow-up mean values were calculated for only participants with complete primary care data.*

##### Table S3. Disaggregated costs

| **Resource items** | **Ibuprofen N=324** | | | **Placebo N=322** | | | **Bootstrap difference (95% CI)** | | |
| --- | --- | --- | --- | --- | --- | --- | --- | --- | --- |
|  | **n** | **Mean** | **SD/SE** | **n** | **Mean** | **SD/SE** | **Adjusted Mean difference** | **Lower CI** | **Higher CI** |
| **Trial medication** | | | | | | | | | |
| Average number of doses received | 324 | 270 | 53 | 322 | 0 | 0 | 271 | 265 | 276 |
| ***Secondary binary short-term outcomes (Mean/SE)*** | | | | | | | | | |
| Sepsis treatment | 306 | 688 | 1857 | 303 | 676 | 1843 | 7.91 | -283 | 299 |
| Coagulopathy | 320 | 13 | 39 | 320 | 11 | 36 | 1.4 | -4 | 7 |
| IVH_1_2 | 324 | 367 | 583 | 322 | 393 | 595 | -30 | -119 | 60 |
| IVH_3_4 | 324 | 233 | 582 | 322 | 177 | 517 | 54 | -30 | 139 |
| Hydrocephalus | 324 | 136 | 665 | 322 | 95 | 559 | 42 | -50 | 134 |
| Cystic PVL | 324 | 150 | 683 | 322 | 91 | 536 | 58 | -34 | 150 |
| Non-cystic PVL | 324 | 10 | 123 | 322 | 10 | 123 | 0.28 | -19 | 20 |
| Gastrointestinal bleeding | 324 | 48 | 287 | 322 | 49 | 287 | -0.4 | -45 | 44 |
| Intestinal perforation | 25 | 9446 | 3560 | 23 | 10267 | 2238 | -607 | -2166 | 952 |
| Pneumothorax | 313 | 31 | 275 | 310 | 29 | 309 | -9 | -53 | 36 |
| Seizure | 313 | 18 | 122 | 311 | 13 | 104 | 5 | -13 | 23 |
| PDA medical treatment | 324 | 169 | 433 | 322 | 325 | 556 | -151 | -228 | -74 |
| PDA Surgical treatment | 324 | 75 | 442 | 322 | 258 | 793 | -186 | -281 | -90 |
| ROP screening | 324 | 212 | 136 | 322 | 225 | 167 | -13 | -37 | 11 |
| ROP treatment | 324 | 196 | 489 | 322 | 197 | 490 | -5 | -76 | 65 |
| Steroids | 323 | 6 | 11 | 322 | 6 | 10 | 0.151 | -1 | 2 |
| Pulmonary haemorrhage | 322 | 144 | 510 | 322 | 108 | 446 | 36 | -36 | 108 |
| Pulmonary hypertension | 324 | 47 | 201 | 321 | 45 | 196 | 1.8 | -28 | 31 |
| NEC requiring surgery | 323 | 316 | 1264 | 322 | 416 | 1438 | -107 | -313 | 97 |
| ***Secondary short-term outcomes (Mean/SD)*** | | | | | | | | | |
| Blood transfusions | 190 | 302 | 140 | 179 | 309 | 143 | -6 | -33 | 22 |
| Exchange blood transfusion | 0 | - | - | 1 | 1301 | - | - | - | - |
| Cerebral ultrasounds | 324 | 495 | 319 | 322 | 489 | 273 | 5 | -38 | 47 |
| ECHO | 309 | 396 | 248 | 307 | 440 | 249 | -41 | -78 | -5.4 |
| Invasive ventilation (endotracheal intubation) | 324 | 3870 | 967 | 322 | 3971 | 748 | -108 | -238 | 22 |
| Non-invasive (NCPAP, HHFNC, nasal ventilation or low flow oxygen) | 324 | 2022 | 705 | 322 | 2133 | 535 | -104 | -197 | -11 |
| Low flow oxygen | 324 | 975 | 513 | 322 | 1004 | 491 | -24 | -102 | 54 |
| Neonatal intensive care days | 313 | 50764 | 38583 | 311 | 52898 | 50508 | -2101 | -8415 | 4213 |
| Neonatal high dependency care days | 307 | 47978 | 37731 | 309 | 50564 | 37252 | -2345 | -8126 | 3435 |
| Neonatal special care | 307 | 16455 | 11621 | 307 | 17488 | 12697 | -953 | -2886 | 981 |
| Days of Inotropic support (hypotension) | 133 | 29 | 31 | 120 | 23 | 25 | 5 | -1 | 11 |
| **Long-term outcomes at 2 years (Mean/SD)** | | | | | | | | | |
| PDA Operation | 150 | -17 | 1018 | 164 | 47 | 785 | -79 | -294 | 136 |
| In Night admission | 74 | 6015 | 7235 | 84 | 6810 | 9857 | -997 | -3572 | 1579 |
| ITU | 8 | 15343 | 13369 | 16 | 19178 | 18240 | -6718 | -26063 | 12627 |
| Visit to the Emergency | 46 | 438 | 432 | 70 | 588 | 742 | -133 | -357 | 92 |
| Auditory visit | 44 | 936 | 1401 | 43 | 737 | 814 | 9 | -375 | 393 |
| Eye visit | 66 | 504 | 504 | 72 | 459 | 441 | 48 | -107 | 202 |
| General medicine visit | 24 | 566 | 253 | 26 | 889 | 1431 | -110 | -586 | 365 |
| Paediatrics/Neonatal follow-up | 96 | 1338 | 925 | 115 | 1604 | 1777 | -235 | -613 | 144 |
| Community paediatrician visit | 30 | 3389 | 9710 | 22 | 1546 | 1014 | 2292 | -2372 | 6955 |
| Other hospital visit | 34 | 1004 | 992 | 56 | 1321 | 1428 | 325 | -848 | 197 |
| Community nurse visit | 55 | 611 | 1015 | 71 | 434 | 538 | 194 | -136 | 523 |
| Dietician visit | 46 | 330 | 331 | 57 | 397 | 443 | -71 | -236 | 93 |
| Physiotherapist visit | 64 | 1191 | 2295 | 66 | 1119 | 1851 | 199 | -558 | 957 |
| Speech therapist visit | 38 | 637 | 889 | 38 | 722 | 956 | -120 | -559 | 319 |
| Other professional visit | 19 | 788 | 721 | 16 | 1999 | 3627 | -157 | -1382 | 1068 |

*The follow-up mean values were calculated for only participants with complete primary care data.*

##### Table S4. Mean total costs

| **Cost** | **Ibuprofen N=324** | **Placebo N=322** | **Bootstrap difference (95% CI)** | | |
| --- | --- | --- | --- | --- | --- |
|  | **Mean**  **(SD)** | **Mean**  **(SD)** | **Adjusted mean** | **Lower CI** | **Higher CI** |
| Initial hospital care up to 36 weeks pma or discharge | 126,465 (66878) | 133,260 (76927) | -6,727 | -17,144 | 3,690 |
| Initial discharge up to 2 years of age | 3,401  (9851) | 4,642  (13010) | -1,177 | -2,889 | 534 |
| Total cost from randomisation to 2 years | 129,867  (69202) | 137,903  (81221) | -7,904 | -18,924 | 3,115 |

##### Table S5 Sensitivity analysis

| **Sensitivity analyses** | **Mean costs (£)** | | **Cost difference (£)** | | | **ICER (£)** |
| --- | --- | --- | --- | --- | --- | --- |
|  | **Ibuprofen** | **Placebo** | **Adjusted mean** | **Lower CI** | **Upper CI** |  |
| Complete dose of Ibuprofen | 121,858 | 128,315 | -6,480 | -16,836 | 3,876 | 120,724 |
| Multiple imputation | 143,080 | 151,225 | -12,044 | -24,976 | 887 | 612,879 |
